# Phase-specific Deep Brain Stimulation revisited: effects of stimulation on postural and kinetic tremor

**DOI:** 10.1101/2022.06.16.22276451

**Authors:** Carolina Reis, Shenghong He, Alek Pogosyan, Nikolaos Haliasos, Hu Liang Low, Anjum Misbahuddin, Tipu Aziz, James Fitzgerald, Alexander L. Green, Timothy Denison, Hayriye Cagnan

## Abstract

**Background:** In Essential tremor (ET), involuntary shaking of the upper limbs during isometric muscle contraction closely reflects the patterns of neural activity measured in the thalamus - a key element of the tremorgenic circuit. Phase-specific deep brain stimulation (DBS) builds upon this observation while using accelerometery of the trembling limb to trigger repetitive electrical perturbations to the thalamus and surrounding areas at a specific time within the tremor cycle. This closed-loop strategy has been shown to induce clinically significant postural tremor relief while delivering less than half the energy of conventional DBS.

**Objective:** The main aim of the study was to evaluate treatment efficacy across different contexts and movement states.

**Methods:** We used accelerometery and a digitizing tablet to record the peripheral tremor dynamics of 4 DBS implanted ET patients while alternating stimulation strategies (no stimulation, continuous open-loop and phase-specific) and movement states (intermittent posture holding and spiral drawing).

**Results:** In addition to observing a suppressive effect of phase-specific DBS on both postural and kinetic tremor, our results reinforce the key role of phase-specificity to achieve tremor control in postural motor states and highlight the difficulty of quantifying phase-dependent effects during continuous movement. Moreover, this study supports the hypothesis that ET patients with more stable tremor characteristics benefit the most from phase-specific DBS.

**Conclusions:** By creating a better understanding of the dynamic relationship between central and peripheral tremor activity, this study provides important insights for the development of effective patient and context-specific therapeutic approaches for ET.

## Introduction

Temporally coordinated neural activity, within and across brain regions, is essential for normal functioning of the brain [1–3]. Disruption of information flow is implicated in many brain disorders and results in hypo- or hyper-synchronised neural activity [4–6]. Therapeutic options that can reverse abnormal brain synchrony in oscillopathies such as Essential Tremor (ET), Parkinson’s disease (PD), epilepsy and obsessive compulsive disorder, range from pharmacological treatments, lesioning, and Deep Brain Stimulation (DBS) [7–12]. However, these therapies often induce widespread effects on the network, which can at times result in side-effects [13–17]. To improve DBS therapy, efforts to create on-demand brain stimulation strategies, i.e., systems that trigger stimulation only when a disease biomarker is sensed, date back to the early 1980s [18].

Among some of the recently developed approaches (see [19] for a review), delivering patterned DBS according to the temporal properties of the pathological neural activity in a closed-loop fashion, is an experimental strategy which has been shown to selectively manipulate synchronisation. By delivering stimulation at a specific timing of the oscillatory neural activity, phase-specific stimulation is capable of either promoting or disrupting neural synchrony [20–26]. In ET, a neurological disease which affects up to 6.3% of the population above 65 years old [27], excessive synchronisation of the cerebellar-thalamo-cortical circuit is thought to drive involuntary shaking of the limbs [28]. Critically, thalamic neural activity closely matches the rhythmic limb acceleration during a postural tremor episode [29,30]. This presents a unique opportunity for guiding stimulation: patient’s tremor can be used to control stimulation and determine the optimal pattern that can modulate tremor. DBS controlled by limb acceleration (phase-specific DBS) was trialled by Cagnan and colleagues [22] and was shown to achieve comparable levels of postural tremor reduction with respect to conventional high-frequency DBS while using less than half of the energy for a subset of the cohort.

Tremor in patients diagnosed with ET emerges not only when a posture is held (i.e., when an isometric contraction occurs; named postural tremor) but also during continuous movement (named, kinetic tremor) [31]. Hence, it is paramount that effects of phase-specific DBS on kinetic tremor are evaluated before this strategy can be considered as a therapeutic option. In line with the above, this study aimed to contrast phase-specific DBS effects in the same patient population during two different tasks. Using the upper-limb tremor of 4 ET patients, we controlled stimulation to the contralateral thalamus or surrounding areas in a closed-loop fashion, while patients were either holding a posture or drawing a spiral. Our results suggest that for a subset of patients: 1) phase-locked stimulation could modulate both postural and kinetic tremor, albeit not to the same degree as conventional high-frequency DBS, and 2) suppressive effects of phase-locked stimulation are maintained as patients alternate between holding a posture and resting their hands.

## Materials and Methods

### Participants

Six patients with ET were recruited for this study. The research project was approved by the local ethics committee in accordance with the Declaration of Helsinki and all patients gave their informed consent to take part in the study. Patients 2 and 5 were excluded from the study; the former due to an incompatibility between the patient’s implanted stimulator (Medtronic Kinetra) and our interface Nexus-D2 investigational device (Medtronic) [32] for phase-locked stimulation, and the latter due to insufficient tremor. On the day of the experiment, a potential diagnosis of dystonic tremor (DT) was raised for patient 3 by the clinical team (Table 1). However, it should be noted that the distinction between ET and DT remains an ongoing topic of discussion.

**Table 1.** Clinical information and stimulation parameters of patients included in the study. ET = Essential Tremor; DT = Dystonic Tremor; PSA = Posterior Subthalamic Area; ViM = Ventral Intermediate Thalamus; Clinical and experimental stimulation parameters: pulse width, amplitude and frequency; NS = No-Stimulation; L = Left; R=Right; B=Battery; HFS = High-Frequency Stimulation; RSS= Random-Search Stimulation; PSS= Phase-Specific Stimulation; N = No; Y=Yes.

| Patients | Diagnose | Meds | Stim target | Clinical parameters | Experimental parameters | Stim electrode and contacts | Experimental conditions | PSS posture | PSS spiral |
| --- | --- | --- | --- | --- | --- | --- | --- | --- | --- |
| 1 | ET | N | PSA | 120µs, 2.5V, 120 Hz | 210µs, 2.5V | (L) 2- B+ | NS/HFS/RSS/NS | N | N |
| 2 | ET/DT | N | PSA | 60µs, 1.1V, 130Hz | 210µs, 1.7V | (R) 9- B+ | NS/RSS/PSS/HFS | Y | Y |
| 3 | ET | Primidone<br>Topiramate | PSA | 60µs, 1.3V, 130Hz | 210µs, 1.3V | (L) 1- B+ | NS/RSS/PSS/HFS | N | Y |
| 4 | ET | N | ViM | 90µs, 3.3V, 130Hz | 210µs, 3.0V | (L) 2- 3+ | HFS/NS/RSS/PSS | Y | Y |

### Recordings

Participants were not asked to interrupt their therapeutic routine. During the recording session, the most tremulous hand was visually assessed and a triaxial accelerometer (Biometrics Ltd, ACL300) was placed on top of the metacarpophalangeal joint of the index finger of this hand. The triaxial signals from the accelerometer sensor were amplified using a Biometrics K800, recorded using a 1401 amplifier (Cambridge Electronic Design) at a sampling rate of 10,417 kHz and downsampled to 1000 Hz for analysis. Tremor was recorded during two distinct motor tasks: posture holding, where patients assumed a tremor provoking posture: folding of the forearms inwards so that both hands were pointed at each other at the level of the nose, and spiral drawing, where inwards and outwards spirals were drawn on a tablet (Sony, Vaio SVF13NA1UM). Displacement of the drawn signal was recorded in the x and y cartesian coordinates (mm) with a varying sampling rate (on average 64Hz ± 2STD), which was up sampled to 100Hz using linear interpolation.

### Experimental Design

The experiment consisted of four stimulation paradigms: 1) No-Stimulation (NS), 2) Random-Search Stimulation (RSS), 3) Phase-Specific Stimulation (PSS) and 4) High-Frequency Stimulation (HFS) conditions.

The NS condition consisted of 2 trials during which the patients were asked to assume a tremor provoking posture for about 1 minute followed by 1 minute of resting and 1 minute of spiral drawing (i.e., 2 baseline postures and 2 baseline spirals). Trial initiation and termination points were derived from the hand movement, by lowpass filtering the triaxial signal (third order Butterworth filter with a cut-off frequency of 2Hz) and visually inspecting patients’ hand position.

Two key parameters were extracted from the NS condition during posture holding: 1) the dominant tremor axis (x, y or z), and 2) the peak tremor frequency, which was kept fixed throughout the experiment. These features were determined from the power spectral densities of the triaxial signals with a minimum frequency resolution of 0.5Hz (Spike 2, Cambridge Electronic Design).

During RSS and PSS, the accelerometer signal from the dominant tremor axis was band-pass filtered between 2-8Hz (Digitimer Neurolog N125/6). This bandpass filtered signal was used to estimate the tremor phase in real-time (based on the instantaneous zero crossings and the average tremor frequency estimated from the posture hold task of the NS condition). For each motor task, once the desired phase of stimulation was detected (detailed below), a TTL pulse was sent out from the 1401 amplifier to a laptop which briefly switched on the implanted stimulator for about 35 ms then switched it off, via an investigational device Nexus-D2 (Medtronic;[32]), closing the loop.

The RSS condition was performed to identify the stimulation phase inducing the largest tremor suppression. To this end, patients alternated between posture holding and spiral drawing approximately eight times (depending on the patient’s level of fatigue), while 12 blocks of 5-second stimulation at a randomly selected phase value (n=12, 0-330 degrees with a 30-degree resolution) were delivered per task (Figure 1).

**Figure 1.**
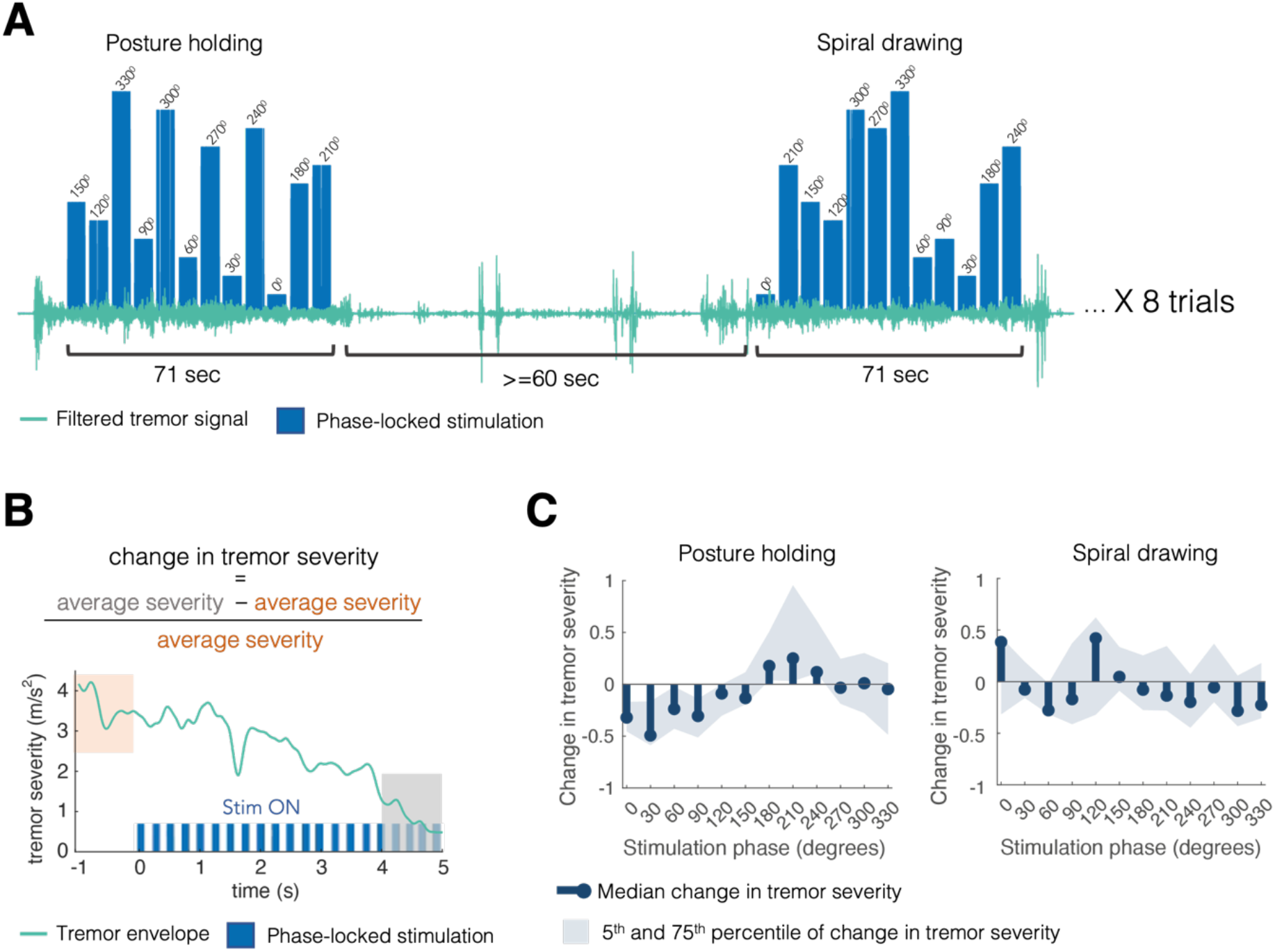
Finding the optimal phase of stimulation in a patient and task-specific manner. Panel A exemplifies how stimulation during the random-search condition was delivered. Stimulation was locked to a certain phase in the tremor cycle (0 - 330 degrees indicated by the blue bars). A stimulation trial lasted 71 seconds where stimulation was locked to 12 different phases for 5 seconds with one second interval in between. The order of stimulated phases was randomised at each trial. Stimulation trials alternated between posture holding and spiral drawing with an inter-trial interval of at least one minute. Up to 8 trials of random-search stimulation were delivered for each motor task. Panel B shows how random-search simulation effects were quantified. The signal from the main tremor axis was band-pass filtered around the peak tremor frequency (± 2Hz) and the instantaneous tremor envelope (green line, tremor severity) was calculated using the Hilbert transform. Impact of stimulation on tremor severity was recapitulated by calculating the change in tremor severity: the average tremor severity at the last second of each 5-second stimulation block (grey shade, 4–5 seconds) normalised with respect to the average tremor severity 1 second prior to the onset of the same stimulation block (orange shade). Change in tremor severity therefore varied between -1 and a positive number: -1 representing complete suppression, 0 indicating no change and a positive number indicating amplification. To find the phase at which stimulation induced the largest tremor suppression, phase-specific effects were summarised using amplitude response curves. Exemplary amplitude response curves for posture and spiral drawing are shown in panel C, where blue stem plots show the median change in tremor severity across different stimulation trials (n=8) at a given phase (n=12). In this specific example, the optimal stimulation phase during posture holding would consist of 30º and for spiral drawing, of 60º.

The PSS condition consisted of six trials; three where stimulation was delivered continuously at the patient-specific phase that yielded the largest tremor suppression in the posture RSS condition, and where patients were asked to hold the tremor provoking posture and interrupt it twice by slowly tapping their hands on their laps and return to posture hold - referred from here onwards as intermittent posture. Tapping occurred approximately at 20 seconds and 40 seconds of stimulation and lasted on average 1300ms±800 SD (ranging from 500 to 2800 msec); and other three trials where patients were asked to draw a spiral for approximately one minute continuously (84 sec±18 SD) while stimulation was delivered at the patient-specific phase affording maximum suppression during spiral drawing in the RSS condition. Due to fatigue, PSS during intermittent posture was only performed by half of the cohort (patients 2 and 4) and spiral drawing was performed by all but patient 1 (cf. Table 1)

### Dominant and Tracked Tremor Axis

The dominant tremor axis derived from the NS posture hold was the one tracked for online-phase estimation during the RSS condition during both motor tasks. During the PSS condition, the tracked axis was re-assigned to the one showing a higher count of stimulation blocks at which the tremor envelope was higher than the other envelopes during RSS posture hold and spiral drawing, respectively (Figure 2B and Figure 3B).

**Figure 2.**
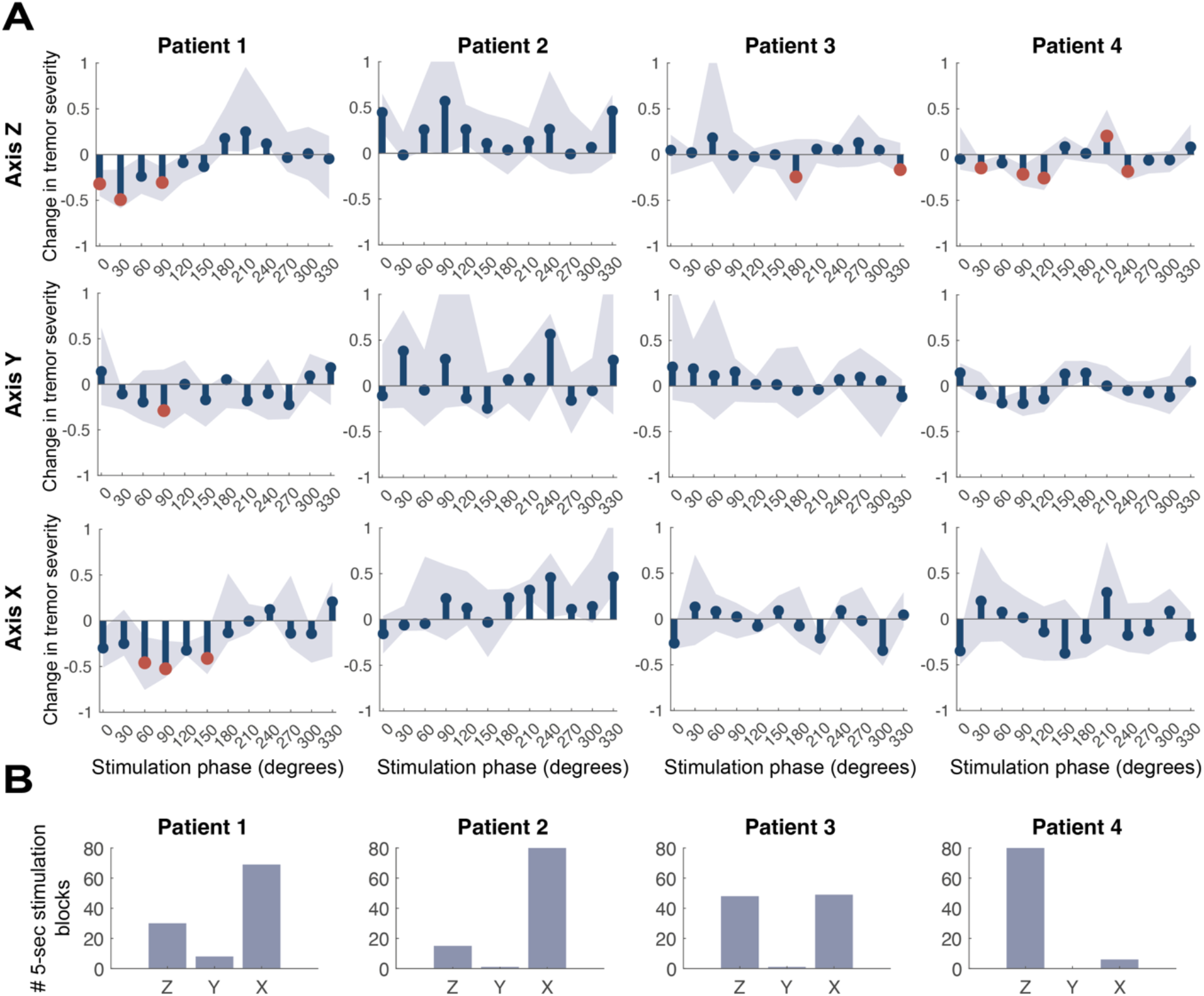
Effects of Random-Search Stimulation on postural tremor amplitude. Blue stem plots in panel A show the amplitude response curves (ARCs) computed for each individual (columns) and each tremor axis (rows) during the posture hold task. ARCs summarise the median change in tremor severity at each stimulation phase, where 0 denotes no modulation of tremor severity, -1 indicates complete tremor suppression and positive values indicate tremor amplification. The shaded light blue area represents the 25th and 75th percentile of change in tremor severity (across RSS trials). Red dots indicate stimulation phases that yielded a significant modulation of tremor severity with respect to that found in the NS condition (α=0.0021, after Bonferroni correction for multiple comparisons). The histograms on panel B, show the count of stimulation blocks at which a given tremor axis had a larger tremor envelope than the remaining 2 axes.

**Figure 3.**
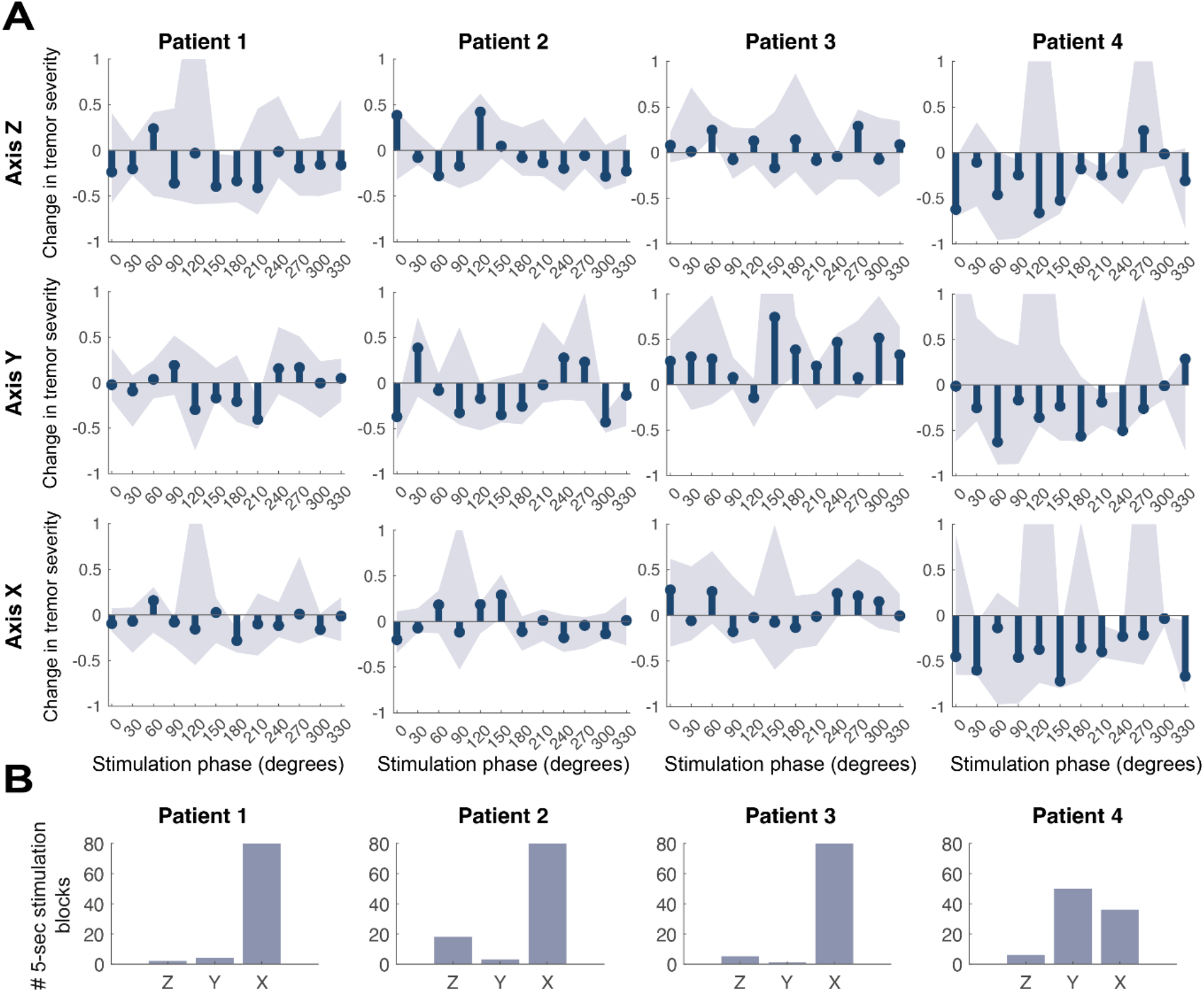
Effects of Random-Search Stimulation on kinetic tremor amplitude. Blue stem plots in panel A show the amplitude response curves (ARCs) computed for each individual (columns) and each tremor axis (rows) during the spiral drawing task. ARCs summarise the median change in tremor severity at each stimulation phase, where 0 denotes no modulation of tremor severity, -1 indicates complete tremor suppression and positive values indicate tremor amplification. The shaded light blue area represents the 25th and 75th percentile of change in tremor severity (across trials, n=8). Red dots indicate stimulation phases that yielded a significantly larger modulation in tremor severity with respect to the no stimulation condition (α=0.0021, after Bonferroni correction for multiple comparisons). The histograms on panel B, show the count of stimulation blocks at which a given tremor axis had a larger tremor envelope than the remaining 2 axes.

### DBS Stimulation Target

DBS electrodes were implanted targeting either the Ventral Intermediate thalamus (ViM) or the fibre tracks immediately inferior to the thalamus known as the Posterior Subthalamic Area (PSA), depending on the hospital site patients had their surgery performed. We have not separated phase-specific effects according to surgical planning (please see Limitation & future directions).

## Data Processing

### Quantification of change in tremor severity during random-search stimulation

Two amplitude response curves (ARCs) were calculated per patient to summarise the median change in tremor severity during each motor task while stimulation was delivered at 12 different phase values. For a detailed description of the analysis pipeline, please refer to Figure 1. To determine whether modulation of postural and kinetic tremor severity during RSS was larger than the natural variability of tremor, first the envelope of the accelerometer signal from the NS condition was derived using the Hilbert transform. Next, 50,000 5-second long segments were randomly selected and the change in tremor severity was calculated. From this distribution, x values were randomly selected and their median calculated (x corresponding to the average number of stimulation blocks delivered at each phase for a given patient, to closely match the median change in tremor severity that created ARCs in the RSS condition). This was repeated 1,000,000 times leading to a surrogate distribution of spontaneous changes in tremor severity. Stimulation effects summarised by ARCs were then compared to the z-scores of this distribution after controlling for multiple comparisons using the Bonferroni method (n=12 phases, α=0.0021).

### Quantification of change in tremor severity while intermittently holding a posture during phase-specific stimulation

The impact of PSS on intermittent postural tremor (posture sporadically interrupted by brief periods of rest where patients slowly tapped their hands on their lap) was evaluated by describing how tremor modulation fluctuated around tapping periods (Figure 4). The delineation of the onset and offset of tapping was achieved visually by inspecting patients’ hand position from the change in DC offset of the low pass filtered triaxial signals (third order Butterworth filter and cut-off frequency of 2Hz).

**Figure 4.**
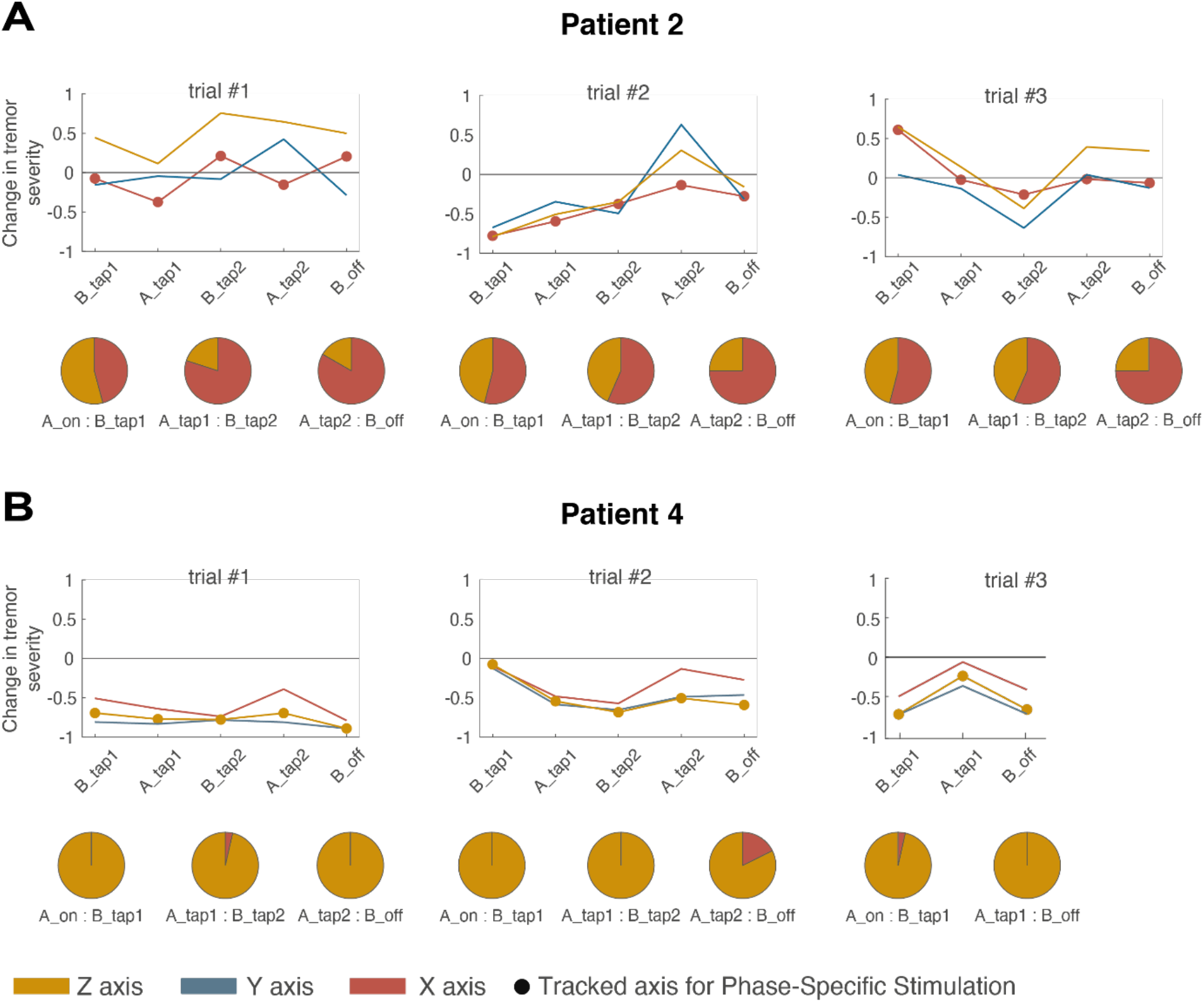
Consistency of stimulation effects are linked to stable contributions of independent oscillators and our ability to identify an effective stimulation phase. The top row of each panel summarises the fluctuations in postural tremor severity (envelope of the band-passed filtered accelerometer signal) for all the three axes at different instances of the intermittent posture task – before tapping the first time (B_tap1); after tapping the first time (A_tap1); before tapping the second time (B_tap2); after tapping the second time (A_tap2); and before stimulation is turned off (B_off). Change in tremor severity consists of a normalised measure of the averaged tremor severity (over 3 seconds) at these five different instances of the task with regards to the average tremor severity before stimulation onset; a change in tremor severity with a positive value indicates tremor amplification, and a negative value indicates tremor suppression. Lines with dots denote the tremor axis being tracked to deliver phase-specific stimulation. The pie charts indicate dominant tremor axis as a ratio for a given epoch (e.g., A_tap1: B_tap2 indicates the epoch from after tapping the first time to before tapping the second time). The label A_on, indicates the time point after which stimulation was turned on.

### Quantification of change in tremor severity while spiral drawing during phase-specific stimulation

Unlike posture holding, spiral drawing emerges from voluntary contributions which continuously evolve over time. Therefore, we have: 1) analysed the accelerometer signal as a composite signal 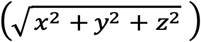, instead of examining a single (dominant) axis, and 2) measured stimulation effects by comparing the average tremor signal envelope across individual spirals drawn during the NS, PSS and HFS conditions, instead of computing changes in the stimulated spiral with respect to tremor prior to stimulation onset. Prior to band-pass filtering and Hilbert transforming the signal to obtain the tremor envelope, the composite tremor signal was concatenated across stimulation conditions and subsequently z-transformed.

Finally, we were interested in exploring whether changes in tremor across stimulation conditions depended on spatial tremor dynamics. To this end, we have processed the digitised spiral signal 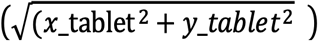 as described above and compared the average power of displacement at tremor frequencies found in polar quadrants (90º to 0º; 0º to -90º; -90º to -180º and -180º to 90º) of each spiral drawn at each condition, using a repeated measures ANOVA.

## Data availability and sharing

The data that support the findings of this study are available from the corresponding author, upon reasonable request.

## Results

With this study we aimed to explore the robustness of phase-specific DBS during different motor states. While phasic-stimulation has proven to be efficient in inducing clinically relevant symptom relief by disrupting critical temporal relationships that are established during postural tremor in ET [22], it remains unknown 1) if stimulation effects during postural tremor are maintained when posture is interrupted by short periods of rest and 2) whether phase-dependent suppressive effects can also be observed during kinetic tremor. By providing a thorough assessment of the spatial and temporal dynamics of tremor while stimulation was delivered at specific instances of tremor, we sought for a better understanding of the key features underlying phase-specific stimulation efficacy.

### Effects of Random-Search Stimulation

To determine the consistency of stimulation’s modulatory effects at each stimulation phase and find the optimal phase of stimulation, median changes in tremor severity for each patient during each motor task, axis and stimulated phase have been contrasted with respect to the corresponding surrogate distributions which summarised the spontaneous (unstimulated) changes in tremor severity.

We found that during posture hold, all ET patients, except for patient two (for whom a possible DT diagnosis has been raised during the recording session), had at least one instance at which stimulation afforded significant tremor modulation (marked as red dots on Figure 2A). It is also worth highlighting the bidirectionality of stimulation effects in patient 4 (Axis Z), where significant suppressive and amplifying effects were observed depending on the specific phase at which stimulation was delivered. During spiral drawing however, none of the 4 patients showed statistically significant phase-dependent stimulation effects on tremor severity (Figure 3A).

### Effects of phase-specific stimulation during intermittent posture

Prolonged stimulation at the optimal phase for postural tremor suppression was delivered to patients 2 and 4 while they intermittently held a posture. Intermittent posture holding meant that the posture sustained during stimulation was sporadically interrupted by brief periods of rest when patients slowly tapped their hands on their lap. Tremor in ET is predominantly an action tremor, meaning that during rest, the involuntary shaking of patients’ limbs stops. Because our paradigm relies on the presence of tremor to deliver stimulation, it is implied that during episodes of rest (tapping periods), the signal used to control stimulation is interrupted.

PSS during intermittent posture was delivered at 0 degrees of the X axis for patient two and at 120 degrees of the Z axis for patient four and yielded very distinct results. Patient 2 showed inconsistent stimulation effects (an average 14% change in tremor severity ± 35% SD) and frequent shifts in the dominant tremor axis from axis Z to axis X (67% in axis X and 34% in axis Z across all trials, Figure 4A). It should be noted that delivering stimulation at 0 degrees did not yield a statistically significant modulation during the RSS condition as well (Figure 2A), indicating that inconsistent results during the PSS condition is mostly driven by the fact that an effective stimulation phase could not be identified for this patient. Conversely, in patient 4, PSS induced a continued suppression of postural tremor (−74% change in tremor severity ±14SD) and the dominant tremor axis was maintained throughout the task (97% in axis Z and 3% in axis X across trials, Figure 4B).

### Effects of prolonged phase-specific stimulation during spiral drawing

Prolonged stimulation at the phase for highest kinetic tremor suppression (despite not statically significant, cf. with Figure 3A) was delivered while patients repeatedly drew spirals inwards and outwards. We found that there was a significant effect of stimulation over kinetic tremor severity at the group level (n=3; repeated measures ANOVA, p<0.001, for both accelerometer and tablet data, Figure 5A). Crucially, despite inducing significant tremor suppression (compared to NS, p<0.006 for accelerometer data and p<0.001 for tablet data), PSS effects on kinetic tremor were less than those of HFS (p=0.006 for accelerometer data and p<0.001 for tablet data).

**Figure 5.**
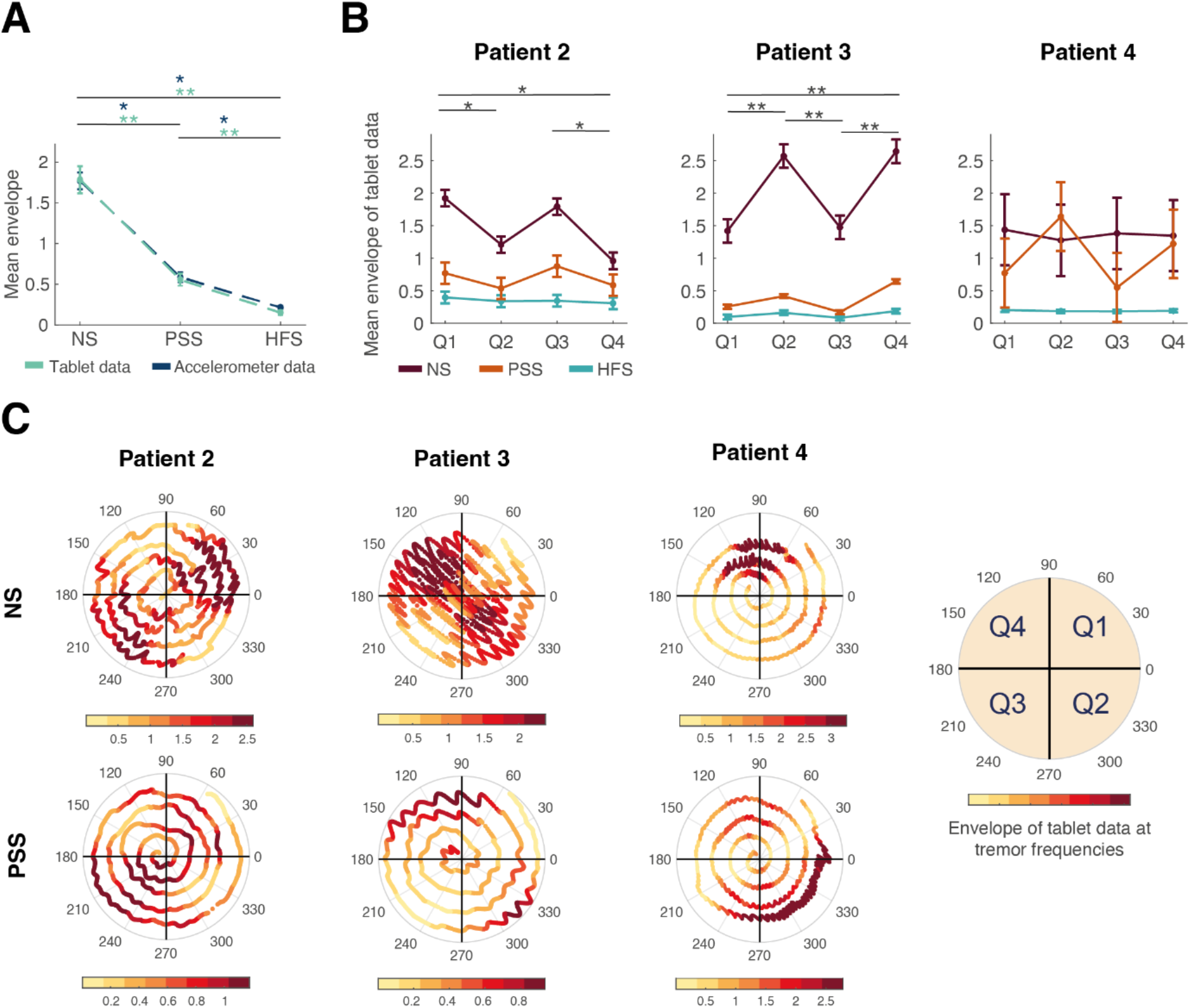
Power and spatial dynamics of kinetic tremor across stimulation conditions. Panel A shows the mean envelope of tablet data (green) and accelerometer data (blue) filtered at tremor frequencies across patients (n=3). Error bars represent ± 1 STD. There was a significant effect of stimulation condition over the mean envelope of tremor at the group level (repeated measures ANOVA, p<0.001 for both tablet and accelerometer data). Mean envelope of data at tremor frequencies was smaller in PSS and HFS when compared to NS (p<0.006* for accelerometer data and p<0.001** for tablet data), and smaller in HFS when compared to PSS (p=0.006* for accelerometer data and p<0.001** for tablet data). Panel B shows the mean ± 1 STD (dots and error bars) of tablet data at tremor frequencies for polar quadrants (Q1, Q2, Q3, Q4) of spirals drawn by individual patients during different stimulation conditions (NS in brown, PSS in orange, HFS in teal). Only patients 2 and 3 show significant effects of stimulation over power of displacement at tremor frequencies as a function of polar quadrants (p= 0.029 and p<0.001 respectively). The consistency of tremor’s spatial manifestation across NS and PSS are quantified in panel B (*p<0.05 and **p<0.001) and depicted in panel C where an exemplary digitised spiral drawn by each patient at NS and PSS conditions are shown. Limits of each spiral’s inset colour bar are set as the minimum and maximum power of displacement at tremor frequencies found across trials of each condition.

At the individual level we found a main effect of stimulation condition over the power of displacement at tremor frequencies (tablet data) for patients 2 and 3 (p<0.001) but not for patient 4 (p=073). Specifically, the level of reduction in tremor-related displacement between NS and PSS conditions was only significant for patients 2 and 3 (table 2). It is worth noting how in the specific case of patient 2, where no significant phase-dependent tremor modulation was found in the RSS conditions of both motor tasks, locking stimulation to the most suppressive phase of the respective motor task was sufficient to induce tremor suppression during spiral drawing in the PSS condition but not to induce sustained postural tremor reduction during intermittent posture holding. Finally, both patients 2 and 3 show an interaction between stimulation effects and spiral polar quadrants (repeated measures ANOVA, p= 0.029 and p<0.001 respectively) and a consistent spatial pattern of tremor across NS and PSS conditions – this is, consecutive quadrants displaying significantly different envelope values (Figure 5B; pairwise comparison, adjusted for multiple comparison with Bonferroni correction: patient 2, p<0.05, Q1/Q2, Q1/Q4, Q3/Q4 and patient 3, p<0.001, Q1/Q2, Q1/Q4, Q2/Q3, Q3/Q4).

**Table 2.** Kinetic tremor reduction during PSS. Mean reduction of tremor-related activity in the spirals drawn by patients 2, 3 and 4. Note that reduction of kinetic tremor for patient 4 during PSS was not significant (Pairwise comparison Bonferroni corrected for multiple comparisons).

| **Patients** | **NS-PSS**<br>**(Envelope of tablet data**<br>**at tremor frequencies [a.u])** | **P-value** |
| --- | --- | --- |
| **2** | 0.779 | 0.002 |
| **3** | 1.655 | $<0.001$ |
| **4** | 0.315 | 0.724 |

## Discussion

The ultimate goal of closed-loop stimulation approaches is to limit the emergence of stimulation induced side-effects and delay the depletion of the neurostimulator’s battery. To achieve this, experimental closed-loop DBS protocols seek to adjust stimulation delivery and parametrisation in a symptom-responsive manner. In ET, different experimental control algorithms for closed-loop DBS have been successful in supressing tremor – either guided by tremor episodes (detected via accelerometery[22,33] or decoded from thalamic local field potentials[34]); or by voluntary movements (sensed via EMG of the upper limb [35,36] or decoded from cortical[37] or thalamic local field potentials[34]). The main difference between the stimulation strategy used in this study (phase-specific DBS) and the aforementioned studies lies in the pattern of stimulation during stimulation periods. The former stimulates at high-frequencies and is therefore agnostic to symptom-related neural activity, while phase-specific DBS aims for neuromodulation of tremor-related neural activity by delivering stimulation in a phase specific fashion. Since to date phase-specific DBS has only been trialled during postural tremor of a single cohort of ET patients, this study has sought to further evaluate the effects of phase-specific DBS on tremor. To do so, we have tested the consistency of phase-dependent effects during intermittent posture and spiral drawing in 4 patients. We found that for selected patients with stable tremor characteristics, continued tremor suppression can be achieved by phase-locked stimulation despite brief interruptions in posture and stimulation. In addition, our results suggest that kinetic tremor during complex motor tasks such as spiral drawing can also be reduced via phase-specific DBS, however not to the same level as HFS.

### Postural tremor reduction with Phase-Specific Stimulation

Our results corroborate the findings reported by Cagnan et al., 2017 and show significant phase-dependent tremor suppression in three out of four patients (Figure 2A). Results from prolonged stimulation at the optimal phase for postural tremor suppression suggest that when tremor is manifested almost exclusively at a single axis, suppressive phase-dependent effects can be maintained during intermittent tremor states (tested here via intermittent posture holding). As our tapping periods were maintained for relatively short periods (1300ms±800 SD), an interesting extension of this paradigm would consist of evaluating stimulation efficacy as a function of tapping period and ascertain whether modulatory effects are maintained regardless of the resting period.

One potential mechanism underlying sustained tremor suppression could be linked to plastic effects similar to those observed when phase-dependent stimulation was delivered according to cortical beta rhythms[24]. Alternatively, sustained tremor suppression during intermittent posture might have resulted from a random perturbation of the tremor network. In our system, stimulation is triggered as soon as the control signal is detected i.e., when both tremor amplitude and phase-tracking efficacy are expected to be very low. At this stage, phase-specific stimulation is thought to be equivalent to random periodic stimulation, which if delivered before tremor can be fully established, might be sufficient to avoid a large tremor rebound. As previously suggested by others [22], it would be interesting to assess whether maintaining stimulation at the optimal phase for suppression continuously (i.e., even in the absence of tremor), would be sufficient to prevent a tremor rebound at the onset of postural and goal-directed movements.

### Kinetic tremor reduction with Phase-Specific Stimulation

This study explores for the first time the impact of phase-specific DBS on ET patients drawing spirals. Our results suggest that delivery of repetitive perturbations to the tremor network phase-locked to the peripheral tremor is capable of supressing kinetic tremor. However, due to complex nature of our motor task and experimental design we were unable to show phase-specificity of this effect. The impact of RSS on tremor was assessed in the same fashion for both posture and spiral so that phase dependent-effects could be compared across different motor states. However, while this method is very well suited to capture the impact of phasic stimulation on postural tremor, during spiral drawing we fall into a sampling problem. Posture emerges from a constant voluntary motor drive, and hence phase-dependent effects of stimulation on tremor can be accurately assessed with instantaneous measurements. Spiral drawing, on the other hand, will have blocks of stimulation at the same phase falling at different instances of the spiral, where different voluntary drives prevail. Considering that tremor and voluntary movement pathways overlap [29,38,39], and that with voluntary movement transient beta oscillations occur across the motor circuit [40,41], movement can be considered as a competing oscillator which can transiently alter tremor output. Hence, the spatial patterning of tremor during spiral drawing (Figure 5B and C) and the design of our search algorithm for the optimal stimulation phase (5-seconds stimulation blocks at 12 different phases in 71 seconds), have introduced noise in our analyses and precluded the identification of effective stimulation phases for tremor modulation (spiral ARCs in Figure 3A).

Nevertheless, by stimulating at the most suppressive phase derived from each individual’s ARC (non-significant), we found a group effect of phase-locked stimulation on tremor envelope. For a nuanced description of spatial tremor dynamics during phase-specific stimulation we have analysed the pen displacement recorded by our tablet. Digitizing tablets have been used in previous studies to detect, characterise and quantify motor dysfunction as well as to aid in the diagnosis of patients with movement disorders [42–46]. This data did not only allow us to quantify the often-observed and reported segregation of tremor in spirals drawn by ET patients when no stimulation is provided [47,48] (Figure 5B and C, NS), but has also shown that such spatial segregation is kept when PSS is effective in reducing tremor (figure 5B and C, PSS).

PSS was capable of reducing tremor significantly in 2 out of 3 patients, but not to the same level achieved by HFS (Figure 5B). It is possible that the phases at which stimulation was delivered, were close to, but not exactly at the individuals’ optimal phase for tremor suppression during spiral drawing. Alternatively, it could be that for spiral drawing, a periodic phase-unspecific perturbation to the tremor network would be enough to dampen but not completely disrupt the central tremorgenic drive for some patients.

### Feasibility of Phase-Specific DBS

Phasic stimulation strategies aim to achieve symptom relief by disrupting the specific phase-alignment that a given brain circuit assumes during a diseased state. In ET, individual fluctuations in diseased states (observed in the periphery as spatial, frequency and/or amplitude changes in tremor) are context-specific, in that they arise from intermittent interactions with competing neural oscillators or altered thalamic afferency from the periphery, cerebellum, cortex, basal ganglia and ascending arousal systems for instance. With the current protocol and methods, under stable circumstances such as holding a posture, phase-dependent effects can be identified and maintained for certain patients. However, in more complex scenarios, such as spiral drawing, where the variability of our control signal is larger, mapping modulatory effects to stimulated phases might prove difficult. In sum, the efficacy of phase-specific DBS relies on how well we can continuously identify the best time to interact with the pathological brain network as different diseased states emerge. Despite being complex and computationally expensive, robust tremor control via phase-specific stimulation might become feasible via future machine learning innovations allowing for a swift sensing of the current network state and appropriate identification of, and adaption to, the optimal phase of stimulation [49,50].

In this cohort, phase-specific stimulation was not as effective as open-loop HFS during both motor states. This raises an important consideration for future applications of this closed-loop stimulation technique: how can we achieve a balance between implementation complexity and therapeutic benefit? Our results and previous work in this field suggest that phase-specific stimulation could be a viable option for a subset of patients with stable tremor characteristics [22,51] and most suited when stimulation side-effects present a limiting factor, enabling us to achieve the appropriate balance between implementation cost and therapeutic benefit. Phasic stimulation also provides a unique opportunity to achieve bidirectional (amplifying and suppressive) effects on neural rhythms and may play a critical role in other neurological and psychiatric conditions where the main aim is to amplify/reinforce certain rhythms.

### Limitations and future directions

Our results were encouraging while showing for the first time how phase-specific stimulation can bring tremor relief during different motor states. Given that only a few cases have been included in this study, it is paramount that the impact of phasic stimulation on kinetic and interrupted postural tremor are validated in a larger cohort. Another limitation of our study is the heterogeneity of DBS surgical targeting - ViM versus PSA (Table 1). Growing evidence indicates that the two targets lead to optimal clinical outcome [52–55], and propose that tremor suppression during stimulation of these (adjacent) targets directly correlates with activation of the same fibre bundles [56–58]. Based on these findings and limited by our sample size, here, ViM phase-specific stimulation effects (n=1) have not been contrasted against those from PSA phase-specific stimulation (n=3).

## Conclusion

Our findings suggest that phase-specific DBS is capable of supressing not only postural but also kinetic tremor in ET patients. While phase-specificity seems to be key to achieve tremor control in postural motor states, periodic phase-unspecific perturbations to the central tremor network seem to be sufficient to yield tremor suppression during continuous movement for a subset of patients. Moreover, this study suggests that for patients with stable tremor characteristics, continued tremor suppression can be achieved.

## Acknowledgements

We would like to thank all our participants for taking part in this study. We would also like to thank Medtronic for supporting the study with the provision of the Nexus-D2 investigational device, and Claudia Sannelli, Duane Bourget, Gaetano Leogrande for their time and support. Finally, we would like to acknowledge Timothy West, Petra Fischer, and Marielle Stam for their help with data collection.

